# Estimating the lives that could be saved by expanded access to weight-loss drugs

**DOI:** 10.1101/2024.06.27.24309551

**Authors:** Abhishek Pandey, Yang Ye, Chad R. Wells, Burton H. Singer, Alison P. Galvani

## Abstract

Obesity is a major public health crisis in the United States (US) affecting 42% of the population, exacerbating a spectrum of other diseases and contributing significantly to morbidity and mortality overall. Recent advances in pharmaceutical interventions, particularly glucagon-like peptide-1 (GLP-1) receptor agonists (e.g., semaglutide, liraglutide) and dual gastric inhibitory polypeptide and glucagon-like peptide-1 (GIP/GLP-1) receptor agonists (e.g., tirzepatide), have shown remarkable efficacy in weight loss. However, limited access to these medications due to high costs and insurance coverage issues restricts their utility in mitigating the obesity epidemic. We quantify the annual mortality burden directly attributable to limited access to these medications in the US. By integrating hazard ratios of mortality across body mass index categories with current obesity prevalence data, combined with willingness to take the medication, observed adherence to and efficacy of the medications, we estimate the impact of making these medications accessible to all those eligible. Specifically, we project that with expanded access, over 43,000 deaths could be averted annually, including more than 12,000 deaths among people with type 2 diabetes. These findings underscore the urgent need to address barriers to access and highlight the transformative public health impact that could be achieved by expanding access to these novel treatments.

## Introduction

Obesity remains a formidable public health crisis in the United States (US), contributing substantially to both morbidity and mortality (1). It is a major risk factor for various chronic diseases, including cardiovascular disease, type 2 diabetes, and myriad forms of cancer. For example, the risk of endometrial cancer in people with severe obesity is seven times higher compared with the rest of the population who are not overweight (2). Among many other causes of mortality and morbidity, obesity is a risk factor for poor medical outcomes from a plethora of infectious diseases, surgical-site infections, and nosocomial infections (3). Compounding the challenge, the obesity prevalence in the US is skewed towards the vulnerable populations, including those with lower socioeconomic status and limited insurance coverage (4). Adults with obesity are likely to spend over $1800 more in medical costs per year than those without obesity (5). Altogether, the economic burden of obesity is immense, with estimated annual medical costs exceeding $170 billion (5).

Historically, the development of pharmaceutical interventions for weight loss has been fraught with challenges, including safety concerns and limited efficacy (6). However, a paradigm shift has occurred with the discovery that drugs originally intended for diabetes management, particularly glucagon-like peptide-1 (GLP-1) and dual gastric inhibitory polypeptide and glucagon-like peptide-1 (GIP/GLP-1) receptor agonists, can induce substantial weight loss (7). These next-generation medications, such as semaglutide, liraglutide, and tirzepatide, have demonstrated remarkable efficacy in clinical trials, resulting in weight reductions previously considered unattainable through pharmaceutical means alone (7). As a result, these medications are increasingly gaining approval from the U.S. Food and Drug Administration (FDA) for the treatment of obesity, ushering in a new era in obesity management (8).

Semaglutide, initially approved in 2017 for type 2 diabetes as Ozempic, expanded its use in 2021 with FDA approval as Wegovy for chronic weight management in individuals aged 12 and older with obesity (Body Mass Index (BMI) ≥ 30 kg/m^2^) or overweight (BMI ≥ 27 kg/m^2^) with at least one weight-related condition, such as hypertension, type 2 diabetes, or high cholesterol. Tirzepatide, another medication in this class, received approval in 2022 for type 2 diabetes as Mounjaro and most recently in November 2023 as Zepbound for chronic weight management in adults with similar criteria to Wegovy.

Despite their promise, widespread access to these medications remains a critical barrier to addressing the escalating obesity epidemic in the US. While semaglutide (Ozempic, Wegovy) and tirzepatide (Mounjaro, Zepbound) have demonstrated efficacy in reducing body weight, their high cost, limited supply, and lack of comprehensive insurance coverage severely limit their accessibility. Without insurance, the monthly cost of these drugs can exceed $1,000, making them unaffordable for many patients (12). This is particularly concerning given that obesity prevalence is higher among economically disadvantaged populations, further entrenching health inequities.

Medicare, the largest US insurer for older adults, does not cover these medications solely for weight loss, impacting many elderly individuals who could benefit from them (9–12). For those on Medicaid, coverage varies widely across states, often requiring patients to meet additional criteria. Additionally, private insurance coverage is inconsistent, especially as many Americans have inadequate insurance with high deductibles and copays they cannot afford. The cost barrier is further compounded by the ongoing need for these medications, as discontinuation often leads to weight regain, necessitating a long-term financial commitment from patients.

In this study, we quantify the potential reduction in annual mortality that could be achieved by expanding access to these innovative obesity medications. Utilizing established correlations between BMI and mortality risk, along with the obesity prevalence data, we estimate the number of lives that could be saved annually if effective weight loss medications were more accessible. The projections underscore the significant public health impact of these novel treatments, informing policy decisions and healthcare practices related to obesity management.

## Results

In the US, more than 40% of adults are classified as obese (BMI ≥ 30; Figure 1A). By applying the hazard ratio for mortality across different BMI categories relative to the normal BMI category (BMI: 18.5–25), we estimate that 48.4% of the total annual deaths in the US occur among individuals categorized as obese (BMI ≥ 30; SI Table 1). According to the current FDA guidelines, more than 45% of the adult US population is eligible for the new weight-loss drugs, encompassing everyone with a BMI of 30 and above, as well as individuals with diabetes with a BMI between 25–30 (Figure 1A, B).

**Figure 1:**
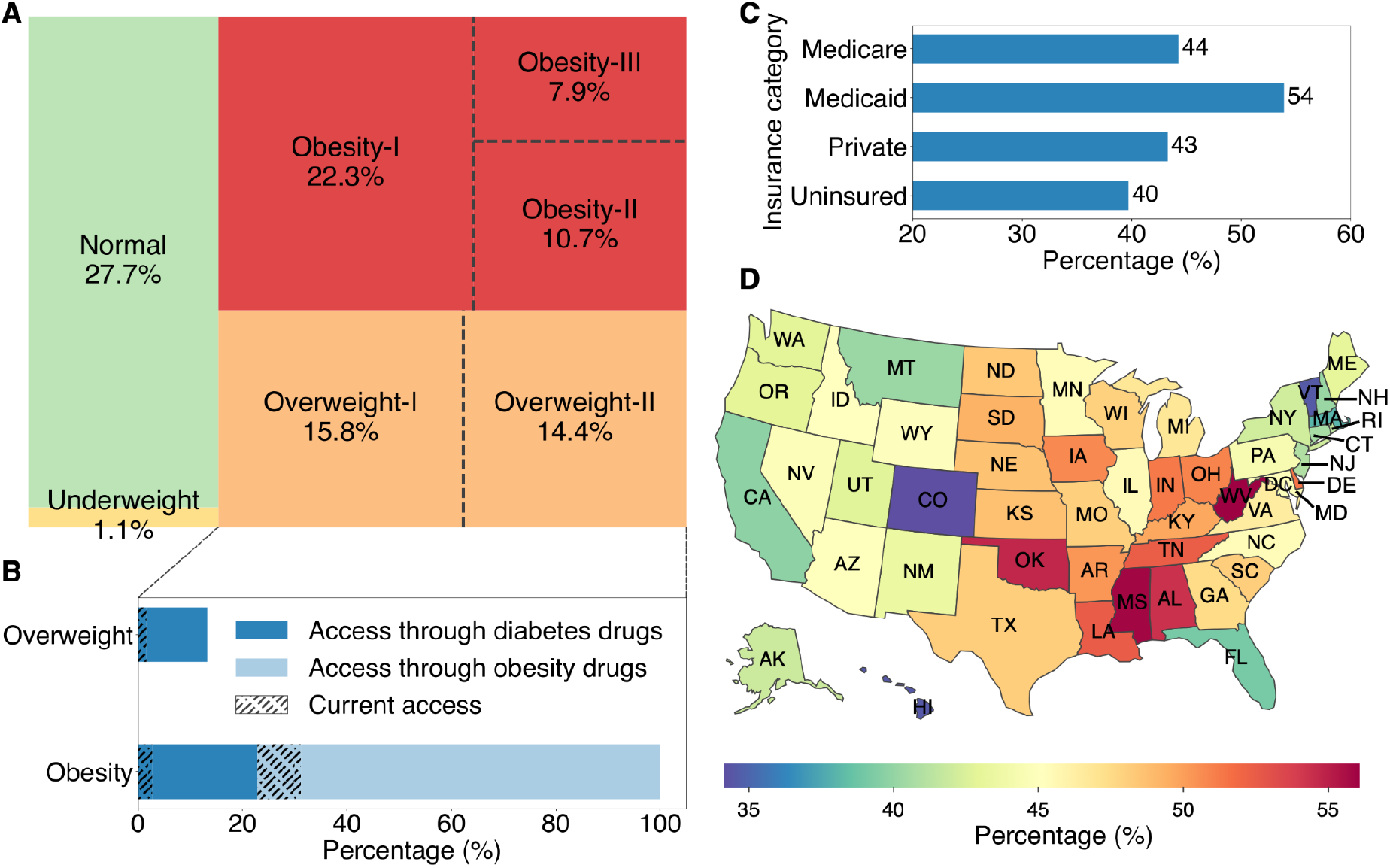
A) Distribution of the US adult population across BMI categories. B) Percentage of population eligible (solid) for weight-loss drugs and proportion that have current access (hatch). C–D) Percentage of eligible population among insurance categories (C) and states (D).

Considering insurance status, 54% of Medicaid recipients and 40% of uninsured individuals are eligible for weight-loss drugs (Figure 1C). State-level variations in obesity and diabetes prevalence result in a range of 34% to 56% of the eligible population, with West Virginia and Mississippi having the highest per capita eligibility (Figure 1D; SI Table 3).

As individuals gain access to these drugs, the distribution of the population by BMI categories is expected to shift towards healthier BMI ranges. Among individuals with diabetes, only 12.2% of those who are overweight (BMI: 25–30) or obese (BMI ≥ 30) on average currently use these weight-loss drugs. Access is even more restricted for obese individuals (BMI ≥ 30) without type 2 diabetes, among whom the current uptake is only 10.8%. Accounting for the adherence rate of people who take obesity drugs (27.2%) and diabetes drugs (48.9%), as well as the eligible individual’s willingness to take the drug, we project how the US population’s BMI distribution would shift. While willingness to take the drugs is implicit in the current uptake rate, our calculations regarding expanded access incorporate the willingness of 75% assessed in survey studies (Methods). Projecting on the basis of current uptake, the change in population distribution by BMI categories is expected to be only marginal (Figure 2), with approximately 1.8% of obese individuals lowering their BMI below 30 (SI Table 4). However, if the access is expanded to all eligible individuals, it would lead to a greater shift in the population distribution by BMI categories, with 12% of obese individuals moving to healthier BMI categories below 30 (SI Table 5). Moreover, 19% of those severely obese (BMI ≥ 40) would reduce their BMI below 40 (SI Table 5). Under a more optimistic scenario of higher willingness to take the drugs (89%) and 100% adherence rate, BMI distribution would shift substantially with 47.1% of obese individuals would have BMI lower than 30 (Figure 2; SI Table 6).

**Figure 2:**
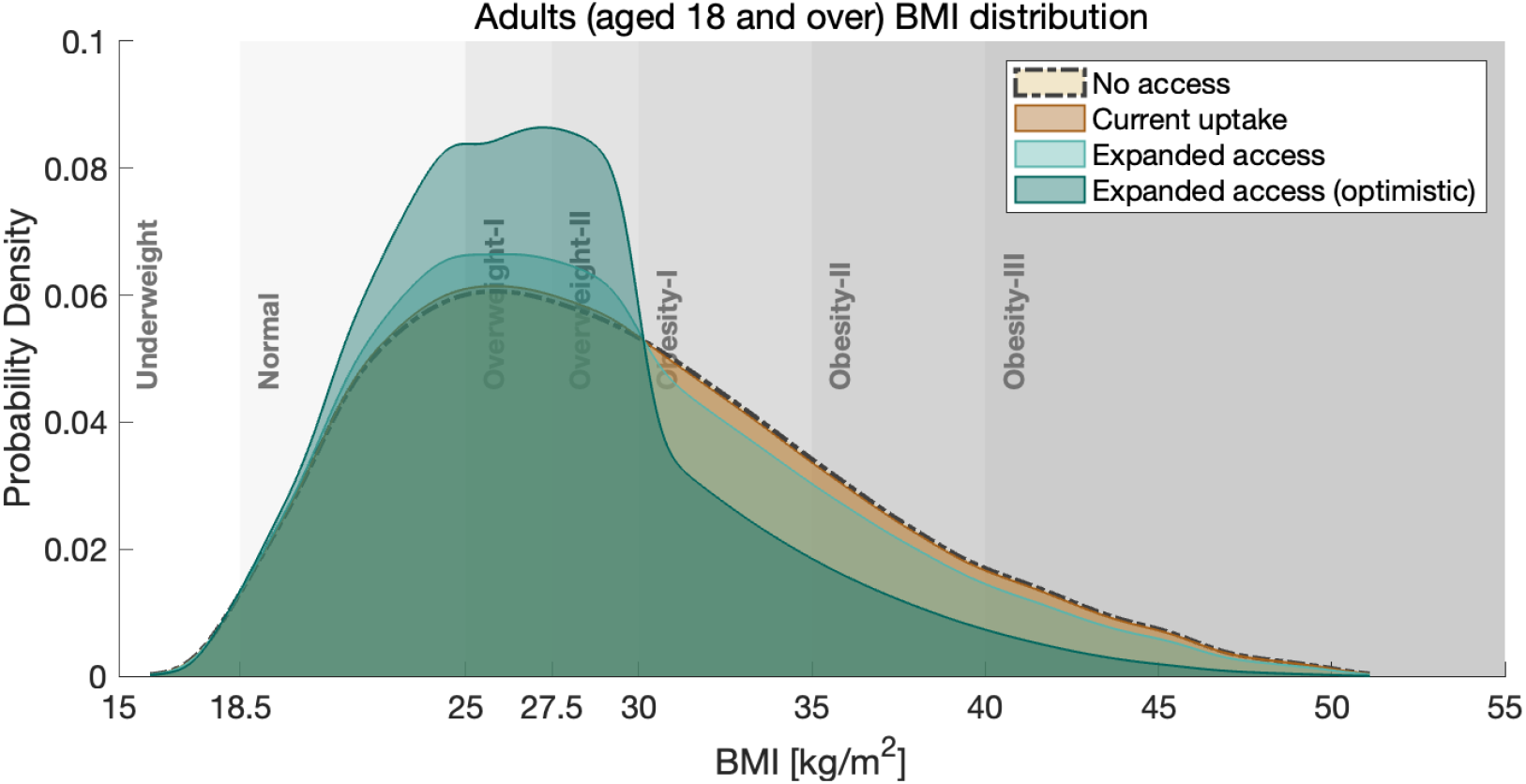
Distribution of the US population across BMI categories with various levels of uptake of weight-loss drugs among eligible individuals.

Based on the projected BMI distributions, we found that at the current uptake level, these weight-loss drugs have the potential to prevent 7,625 deaths annually [95% Uncertainty Interval (UI): 7,613 – 7,636], of which the majority (71%) is expected to be among individuals with private insurance. However, if access were expanded to all eligible individuals, the number of lives saved could rise by 43,719 annually [95% UI: 43,679 – 43,759], of which 12,124 [95% UI: 12,059 – 12,190] would be among overweight and obese individuals with type 2 diabetes (Table 1). With the expanded access, 10,159 [95% UI: 10,139 – 10,178] deaths averted would be among Medicare beneficiaries and 4,245 [95% UI: 4,241 – 4,249] would be among uninsured. Under the optimistic scenario of expanded access, as many as 171,749 [95% UI:171,591 – 171,907] annual deaths could be averted.

**Table 1:** Mortality averted under the current level of access to weight-loss drugs among eligible and 100% access among eligible.

|  |  | Additional lives saved annually |  |
| --- | --- | --- | --- |
|  |  | No access to current uptake | Current to expanded access |
|  |  | Mean [95% UI] | Mean [95% UI] |
| Age group | ≥ 18 | 7,625 [7,613 – 7,636] | 43,719 [43,679 – 43,759] |
|  | 18–64 | 6,884 [6,873 – 6,894] | 33,560 [33,523 – 33,596] |
|  | ≥ 65 | 741 [737 – 744] | 10,159 [10,139 – 10,178] |
| Drug access | Diabetes | 2,332 [2,319 – 2,345] | 12,124 [12,059 – 12,190] |
|  | Obesity | 5,293 [5,282 – 5,304] | 31,595 [31,531 – 31,659] |
| Insurance category | Private | 5,424 [5,416 – 5,433] | 20,329 [20,305 – 20,352] |
|  | Medicaid | 1,459 [1,457 – 1,462] | 8,986 [8,976 – 8,996] |
|  | Medicare | 741 [737 – 744] | 10,159 [10,139 – 10,178] |
|  | Uninsured | 0 [0 – 0] | 4,245 [4,241 – 4,249] |

Considering the geographic distribution of obesity and diabetes on a per capita basis, expanded access to weight-loss drugs among eligibles could lead to an annual mortality reduction of 10 to 16 deaths per 100,000 population. While all states could achieve a reduction of at least 10 deaths per 100,000 population, West Virginia, Mississippi, and Oklahoma are expected to experience the largest per capita reduction (Figure 3). By solely expanding access to obese individuals without type 2 diabetes, more than 40% of the states could still expect a reduction of more than 10 deaths per 100,000 population.

**Figure 3:**
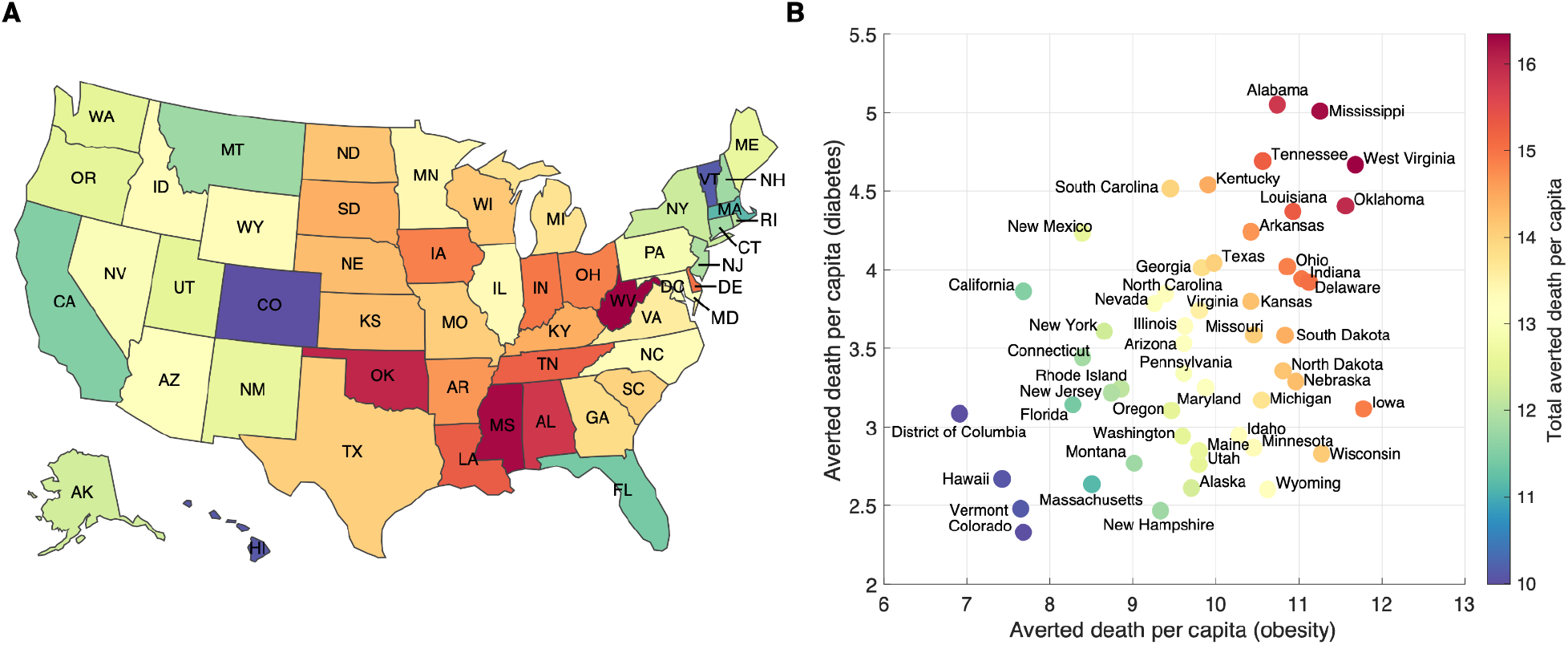
A) State-level annual deaths averted per 100,000 population. B) Distribution of averted deaths per capita among overweight and obese individuals with type 2 diabetes and obese individuals without type 2 diabetes.

## Discussion

As a major risk factor for several chronic diseases, obesity is a national public health crisis. Our findings highlight the immense promise of the new generation of weight-loss drugs to mitigate the mortality and morbidity associated with obesity and diabetes. We estimated that if everyone eligible had access to these weight-loss drugs, the obesity prevalence would decline from 42% (13) to 37%, averting over 51,000 deaths every year. At the current level of uptake, our projections indicate that 7,625 lives would be saved annually, less than 15% of the deaths that could be averted if access were expanded.

Limited access stems from a combination of financial barriers, supply constraints, and restrictive insurance coverage. Although insurance typically covers these medications for diabetes treatment, coverage for weight loss is less consistent, often requiring patients to pay out-of-pocket or face restrictive insurance policies (14). Furthermore, 25.6 million Americans are uninsured (15) and more than 80 million are inadequately insured (16). Compounding the challenges, the high prices of these drugs are significantly more expensive in the US compared to other countries (17). Access challenges have created a market for counterfeit drugs, further complicating access and safety concerns (18). To address the concerns, the US Senate has initiated an investigation regarding the excessive profit margins (19, 20) of these life-changing medications to expand availability to a wider population (21).

Currently those uninsured with diabetes or obesity have no access to these innovative weight-loss drugs, and access is challenging even for those with coverage. For example, despite the widespread need for these medications among elderly aged 65 and above, Medicare Part D only covers the medications for diabetes management or cardiovascular risk reduction (22). Our results show that expanded access to these drugs among elderly with obesity would lead to 10,159 fewer annual deaths among them. Adults under 65 on Medicaid face a higher risk of obesity, with a 27% greater likelihood compared to those with private insurance (23). Accounting for this disproportionate burden, we estimate that if access to these medications were expanded, 27% of the 33,560 annual deaths projected to be averted would occur among Medicaid beneficiaries. However, accessing these medications through Medicaid remains challenging and varies significantly by state.

While Ozempic is ostensibly covered for diabetes treatment under Medicaid in all states, access often involves additional steps, such as prior authorization or trying alternative medications first (28). Coverage for Wegovy, specifically for chronic weight management, is even more restricted under Medicaid (26, 27). Our state-level results demonstrate that high-burden states for obesity and diabetes such as West Virginia, Mississippi, and Oklahoma are likely to avert most per capita deaths by expanding access to all eligible. While Ozempic is covered under Medicaid for type 2 diabetes in these states, they all require prior authorization or step therapy (28–30). Access to Wegovy is similarly restricted, with Mississippi Medicaid not covering it for weight loss alone.

While our results are likely to be robust, several factors may influence them. For example, our estimates are based on available data regarding the effectiveness and utilization of these medications. Specifically, the hazard ratio for mortality stratified by BMI categories used in our calculations was based on a study among white adults in the US (31). However, another retrospective cohort study of US adults of all races found hazard ratios that are consistent with the estimates used for our study (32). Nonetheless, our estimates are conservative in many regards. For example, overweight individuals with a BMI between 27 and 30 are eligible for these weight loss drugs if they have comorbidities such as diabetes, hypertension, or high cholesterol according to FDA guidelines. In our analysis, we conservatively only considered individuals in the overweight-II category with type 2 diabetes as eligible. Also, we used data on semaglutide to parameterize the efficacy of these drug types. Initial data indicates that the most recently approved tirzepdatide may be even more efficacious (33).

Expanding access to obesity medications such as GLP-1 receptor agonists and dual GIP/GLP-1 receptor agonists, and prioritizing obesity as the primary condition for treatment could significantly reduce mortality rates and alleviate the economic burden of obesity-related healthcare costs. The treatment of obesity often leads to improvements in other chronic diseases, including cardiovascular disease and diabetes, as well as diseases that reduce quality of life, such as rheumatoid arthritis (34). Therefore, long-term benefits of improved access could extend beyond mortality reduction, potentially decreasing obesity-related comorbidities with widespread. Even for acute infectious diseases, such as influenza and COVID-19, obesity is a major risk factor for elevated severity and mortality. Our findings provide compelling evidence for the transformative impact that expanded access to these medications could have on improving the public health of the nation. This underscores the urgency of addressing access barriers, including affordability, insurance coverage, and prescriber awareness. Such policies could galvanize a new era of American well-being and prosperity.

## Methods

We first stratified the US population into seven categories based on their BMI: BMI < 18.5, BMI 18.5–25, BMI 25–27.5, BMI 27.5–30, BMI 30–35, BMI 35–40, and BMI ≥ 40 (31). We then expressed the annual mortality in the US as a linear combination of annual deaths in each BMI category, using hazard ratios of mortality for each category compared to the reference category (BMI: 18.5–25) (31). This allowed us to derive the annual mortality rates for individuals in each BMI category.

To assess the impact of new weight-loss drugs, we recalculated the annual mortality in the US by applying the BMI category-specific annual mortality rates to the new population distribution across various BMI categories resulting from the weight loss associated with drug use.

We calculated the new BMI distribution resulting from drug use as follows. Anthropometric reference data from the Centers for Disease Control and Prevention (CDC) (35) provided the age and gender-specific distribution of height, weight, and BMI. Through Monte Carlo sampling of height and weight for each age and gender group, we derived 1000 US population-level samples indexed by weight, height, BMI, age, and gender. Each population sample includes 100,000 people representing the US population stratified across the seven BMI categories.

Next, with each population sample, we modeled the proportion of individuals in each BMI category who would lose weight using diabetes/obesity drugs and calculated their projected BMI. If the new BMI fell into a different BMI category, the individuals were moved accordingly, and the proportion of the total population in each BMI category was recalculated.

The proportion of individuals within a specific BMI category who experience weight loss depends on multiple factors: the proportion of individuals eligible for weight-loss medications, the proportion of eligible willing to take the drugs, the adherence rate among those taking the drug, and the effectiveness of the drugs themselves. These medications can be prescribed for both obesity and type 2 diabetes, with varying eligibility criteria. According to FDA guidelines, obesity medications can be prescribed to individuals with a BMI of 30 or higher, or to those with a BMI between 27 and 30 who have at least one obesity-related condition such as diabetes, hypertension, or high cholesterol. In contrast, all individuals with type 2 diabetes are eligible for diabetes medications and would experience weight loss if their BMI is over 25. Therefore, everyone with a BMI of 30 and above is considered eligible for weight-loss drugs. Of individuals with a BMI between 25 and 30, only those with diabetes are considered eligible in our study.

We used survey studies, prescription data, and cohort studies to parameterize the current uptake rate of the drugs among eligible populations for obesity and for diabetes as 10.8% (36) and 10.7%–13.6% (37, 38), respectively. Informed by Kaiser Family Foundation’s health tracking poll, we took into account that the proportion of eligible individuals’ willingness to use the drugs is 75% (39). In accordance with the clinical trials, we incorporate an adherence rate of 48.9% for individuals with diabetes (40) and 27.2% for individuals taking drugs solely for weight loss (41). Weight-loss drugs are associated with an average weight loss of 16.9% in individuals without type 2 diabetes (42). For individuals with type 2 diabetes, the weight loss is approximately 40% lower than that observed in those without diabetes (43).

We estimated annual deaths averted under two scenarios: the current uptake and expanded access. Assuming that drugs are fully accessible to everyone eligible, uptake under expanded access depends on individual-level decisions to take the drugs. We distributed the national estimates of averted deaths across insurance categories and US states, using the population size, proportion of individuals eligible for weight-loss drugs, and drug accessibility within each group as weights (SI Appendix). The proportion of the eligible population was calculated based on the prevalence of obesity, diabetes, and overweight/obesity among diabetic patients within each group.

For results stratified by insurance category, all adults aged 65 and over are on Medicare, while adults aged 18 to 64 are on Medicaid, private insurance, or are uninsured. Insurance coverage data for adults aged 18 to 64 was obtained from the CDC’s National Health Interview Survey (44). The proportion of individuals eligible for obesity/diabetes drugs was informed by integrating the differential risks across insurance categories with national-level diabetes and obesity prevalence data (23, 25, 35, 45). Under the current access, high-cost weight-loss drugs would be prohibitive for people without insurance, leading to no averted deaths among this cohort. Under the expanded access scenario, we modeled equity of drug accessibility across all insurance categories. For state-level results, we assumed equal drug accessibility across all states for both scenarios. We normalized state-level prevalence of obesity, diabetes prevalence, and overweight/obesity among diabetic patients to align with national-level data (46, 47).

## Data Availability

All publicly available data used for the study along with computational code written in Python for data analysis is publicly available at jianan0099/ObesityInaccessibility

## Acknowledgments

APG acknowledges support from Burnett Endowment and APG, YY, CRW and AP support from the Notsew Orm Sands foundation. The funding source had no influence on the methodology, analysis, or interpretation of the results.

