## Supporting Information for "Estimating the lives that could be saved by expanded access to weight-loss drugs"

**Abhishek Pandey, Yang Ye, Chad R. Wells, Burton H. Singer and Alison P. Galvani (complete author list)**

**Burton H. Singer**
****
**Alison P. Galvani**
****

**This PDF file includes:**

Supporting text
Figs. S1 to S4
Tables S1 to S7
SI References

### Supporting Information Text

All publicly available data used for the study along with computational code written in Python for data analysis is publicly available at (1).

### 1. US population by Body-Mass Index (BMI) category and their hazard ratios for all-cause mortality

Data from a pooled analysis of 19 prospective studies (2), provides gender-specific hazard ratios for all-cause mortality across nine BMI categories (15.0 to 18.4, 18.5 to 19.9, 20.0 to 22.4, 22.5 to 24.9, 25.0 to 27.4, 27.5 to 29.9, 30.0 to 34.9, 35.0 to 39.9, and 40.0 to 49.9) measured in kg/m$^2$. We use this data to estimate overall hazard ratios for seven standard BMI categories: underweight (<18.5), normal (18.5–25.0), overweight-I (25.0–27.5), overweight-II (27.5–30.0), obesity-I (30.0–35.0), obesity-II (35.0–40.0), and obesity-III (≥40.0). The normal BMI category (18.5–25.0) is used as the reference category. The hazard ratio for all-cause mortality in each BMI category is calculated as the ratio of the all-cause mortality rate in that category to the rate in the reference category.

Let $N_F$ and $N_M$ denote the numbers of female and male participants in the pooled analysis, respectively. The numbers of female and male participants in non-standard BMI category $i$ are $N_F f_i^F$ and $N_M f_i^M$, respectively, where $f_i^F$ and $f_i^M$ represent the proportions of participants in non-standard BMI category $i$ among female and male participants, respectively. Note that $\sum_{i=1}^{9} f_i^F = 1$ and $\sum_{i=1}^{9} f_i^M = 1$.

Denote hazard ratios among females and males for non-standard BMI category $i$ as $\lambda_i^F$ and $\lambda_i^M$, respectively. We then calculate the hazard ratio $\lambda_j$ (Table S1) for standard BMI category $j$ using the following formula:

$$\lambda_j = \frac{\left[\sum_{i\in NS(j)} N_F f_i^F \lambda_i^F + N_M f_i^M \lambda_i^M\right] / \left[\sum_{i\in NS(j)} N_F f_i^F + N_M f_i^M\right]}{\left[\sum_{i\in NS(m)} N_F f_i^F \lambda_i^F + N_M f_i^M \lambda_i^M\right] / \left[\sum_{i\in NS(m)} N_F f_i^F + N_M f_i^M\right]}, \quad [1]$$

where $j \in \{1,2,3,4,5,6,7\}$ represents the standard BMI category and $NS(j)$ is the set of non-standard BMI categories that comprise the standard category $j$. In particular, $NS(2) = \{18.5 \text{ to } 19.9, 20.0 \text{ to } 22.4, 22.5 \text{ to } 24.9\}$ represents the normal BMI category (18.5–25.0) and $m = 2$ corresponds to the reference category in the standard categorization (i.e., $\lambda_2 = 1$).

### 2. Weight, height, and BMI distribution in the US population without access to weight-loss drugs

In this study, we focus on US adults aged 18 years and older. Anthropometric reference data from the Centers for Disease Control and Prevention (CDC) (3) provide the means and 5th, 10th, 15th, 25th, 50th, 75th, 85th, 90th, and 95th percentiles of body measures (weight, height, and BMI) for the US population, categorized by gender-age groups. We approximate the weight, height, and BMI distributions for each gender-age group using empirical cumulative distribution functions (ECDFs) with linear interpolation.

For a specific gender-age group (gender $g \in G = \{\text{Male}, \text{Female}\}$ and age group $a \in A = \{18, 19, 20-29, 30-39, 40-49, 50-59, 60-69, 70-79, 80+\}$, we denote the weight, height, and BMI values at the $p_k$th percentile as $w_{p_k}^{g,a}$, $h_{p_k}^{g,a}$, and $b_{p_k}^{g,a}$, respectively. Here, $k \in [1,9]$, $k \in \mathbb{Z}$, $p_{k+1} > p_k$, and $p_k \in \{5, 10, 15, 25, 50, 75, 85, 90, 95\}$.

The ECDF for weight in this gender-age group is defined as:

$$F_W^{g,a}(x_W) = \begin{cases} \max\{0, 0.05 + \frac{x_W - w_5^{g,a}}{w_{10}^{g,a} - w_5^{g,a}} 0.05\}, & x_W < w_5^{g,a}, \\ 0.01 p_k, & x_W = w_{p_k}^{g,a}, \\ 0.01 p_k + (p_{k+1} - p_k) \frac{x_W - w_{p_k}^{g,a}}{w_{p_{k+1}}^{g,a} - w_{p_k}^{g,a}} 0.01 & w_{p_k}^{g,a} < x_W < w_{p_{k+1}}^{g,a}, \\ \min\{1, 0.95 + \frac{x_W - w_{95}^{g,a}}{w_{95}^{g,a} - w_{90}^{g,a}} 0.05\}, & x_W > w_{95}^{g,a}, \end{cases} \quad [2]$$

where $F_W^{g,a}(x_W) \in [0,1]$. Similarly, the ECDFs for height and BMI in this gender-age group are defined as:

$$F_H^{g,a}(x_H) = \begin{cases} \max\{0, 0.05 + \frac{x_H - h_5^{g,a}}{h_{10}^{g,a} - h_5^{g,a}} 0.05\}, & x_H < h_5^{g,a}, \\ 0.01 p_k, & x_H = h_{p_k}^{g,a}, \\ 0.01 p_k + (p_{k+1} - p_k) \frac{x_H - h_{p_k}^{g,a}}{h_{p_{k+1}}^{g,a} - h_{p_k}^{g,a}} 0.01 & h_{p_k}^{g,a} < x_H < h_{p_{k+1}}^{g,a}, \\ \min\{1, 0.95 + \frac{x_H - h_{95}^{g,a}}{h_{95}^{g,a} - h_{90}^{g,a}} 0.05\}, & x_H > h_{95}^{g,a}, \end{cases} \quad [3]$$

and

$$F_B^{g,a}(x_B) = \begin{cases} \max\{0, 0.05 + \frac{x_B - b_5^{g,a}}{b_{10}^{g,a} - b_5^{g,a}} 0.05\}, & x_B < b_5^{g,a}, \\ 0.01 p_k, & x_B = b_{p_k}^{g,a}, \\ 0.01 p_k + (p_{k+1} - p_k) \frac{x_B - b_{p_k}^{g,a}}{b_{p_{k+1}}^{g,a} - b_{p_k}^{g,a}} 0.01 & b_{p_k}^{g,a} < x_B < b_{p_{k+1}}^{g,a}, \\ \min\{1, 0.95 + \frac{x_B - b_{95}^{g,a}}{b_{95}^{g,a} - b_{90}^{g,a}} 0.05\}, & x_B > b_{95}^{g,a}, \end{cases} \quad [4]$$

respectively, where $F_H^{g,a}(x_H) \in [0,1]$ and $F_B^{g,a}(x_B) \in [0,1]$.

We calculate the percentage of the population in BMI category $i$ among individuals in gender group $g$ and age group $a$ as

$$O_i^{g,a} = F_B^{g,a}(\mathcal{B}_i^{max}) - F_B^{g,a}(\mathcal{B}_i^{min}), \quad [5]$$

where $\mathcal{B}_i^{max}$ and $\mathcal{B}_i^{min}$ are the upper and lower bounds of BMI values in category $i$, respectively. Then, the percentage of the population in BMI category $i$ among adults (Table S1) is:

$$O_i = \sum_{g \in G} \sum_{a \in A} p_{g,a} O_i^{g,a}, \quad [6]$$

where $p_{g,a}$ (Table S2) is the proportion of the population in gender group $g$ and age group $a$ among all adults (4). Consequently, the percentage of the population in BMI category $i$ among adults in age group $a$ is $O_i^a = \frac{\sum_g p_{g,a} O_i^{g,a}}{\sum_g p_{g,a}}$. Since CDC data on body measures was not available for $60-64$ and $65-69$ age groups, we assume a uniform distribution for $60-69$ age group. Thus, we calculate the percentage of the population in BMI category $i$ among adults aged 18 to 64 and aged 65 and over as

$$O_i^{18-64} = \frac{\sum_{a \in \{18,19,20-29,30-39,40-49,50-59\}} p_a O_i^a + p_{60-69} O_i^{60-69}/2}{\sum_{a \in \{18,19,20-29,30-39,40-49,50-59\}} p_a + p_{60-69}/2}, \quad [7]$$

and

$$O_i^{65+} = \frac{\sum_{a \in \{70-79,80+\}} p_a O_i^a + p_{60-69} O_i^{60-69}/2}{\sum_{a \in \{70-79,80+\}} p_a + p_{60-69}/2}, \quad [8]$$

respectively, where $p_a = \sum_g p_{g,a}$ is the proportion of adults in age group $a$.

**A. Population-level sample.** While data from the CDC provided distributions of weight, height, and BMI for each gender-age group individually, it did not provide their joint distribution of weight and height with BMI. To address this, we used Monte Carlo sampling to infer the joint distribution by constructing a population-level sample indexed by their height, weight, and corresponding BMI. Specifically, we generated 1000 population-level ensembles representing the US adults indexed by their weight, height, BMI, age, and gender. Each population sample includes 100,000 people (i.e., population-level sample size $N_{pop} = 100,000$), representing the US population stratified across the seven BMI categories. For each specific gender-age group in the data, we sampled heights and weights from their given distributions and used the corresponding BMI distribution as a weight for acceptance criteria. We accepted 1000 samples for each gender-age group (Figs. S2-S4, Table S2).

### 3. Annual deaths without access to weight-loss drugs

Let $P_{total}$ denote the total US population size and $p_{\geq 18}$ the proportion of adults 18 years and older (Table S7), the number of adults is then given by:

$$P = p_{\geq 18} P_{total}. \quad [9]$$

Assuming $\overline{\mu}$ represents the annual mortality rate in the absence of access to any weight-loss drugs, the annual deaths without drug access ($\overline{\mu}P$) can be expressed as the sum of the mortality experienced by individuals in different BMI categories, as

$$\overline{\mu}P = \sum_{i=1}^{n} \mu_i O_i P, \quad [10]$$

where $n = 7$ is the number of BMI categories and $\mu_i$ represents the annual mortality rate without drug access within BMI category $i$, respectively. Since $\mu_i = \lambda_i \mu_2$, we can express total annual deaths as

$$\overline{\mu}P = \mu_2 P \sum_{i=1}^{n} \lambda_i O_i. \quad [11]$$

Thus,

$$\mu_2 = \frac{\overline{\mu}}{\sum_{i=1}^{n} \lambda_i O_i}, \quad [12]$$

and the share of annual mortality by BMI category $i$ is $\mu_i O_i / \overline{\mu}$ (Table S1).

### 4. Individuals eligible for weight-loss drugs

Weight-loss drugs can be prescribed as medications for obesity as well as type 2 diabetes. The eligibility criteria for new obesity drugs, such as tirzepatide (Zepbound) and semaglutide (Wegovy), typically include adults with obesity or overweight who have at least one weight-related condition. Specifically, these drugs are approved for:

- Adults with a BMI of 30 kg/m² or greater.
- Adults with a BMI of 27 kg/m² or greater who also have a weight-related health condition such as type 2 diabetes, high blood pressure, or high cholesterol.

In our study, we only consider type 2 diabetes as an additional criterion for individuals in the overweight-II category. Although many people with a BMI between 27–30 may also have other comorbidities that make them eligible for weight-loss drugs, we conservatively exclude them in our calculations. Also, tirzepatide and semaglutide were previously approved for the treatment of type 2 diabetes under the brand names Mounjaro and Ozempic, respectively. Any adults with type 2 diabetes are eligible for these drugs irrespective of their BMI.

**A. Proportion of individuals in each BMI category eligible for weight-loss drugs.** Based on the eligibility criteria above, everyone with a BMI of 30 and above is considered eligible for weight-loss drugs. While adults with type 2 diabetes are eligible, individuals with a BMI below 25 are less likely to be prescribed weight-loss drugs and if prescribed would be advised to ensure a balanced diet for avoiding weight loss. Therefore, in our analysis, we assume that no one with a BMI under 25 is eligible. Of individuals with a BMI between 25 and 30, only those with type 2 diabetes are eligible. Denote the proportion of individuals eligible for weight-loss drugs in BMI category $i$ as $e_i$, we have

$$e_i = \begin{cases} 0 & \text{if } i \in \{1,2\}, \\ d_i & \text{if } i \in \{3,4\}, \\ 1 & \text{if } i \in \{5,6,7\}. \end{cases}$$

Here, $d_i$ represents type 2 diabetes prevalence among individuals in BMI category $i$. Given that type 2 diabetes accounts for the majority of diabetes cases (5), we assume all diabetes patients are type 2 patients. As patients with diabetes are more likely to be prescribed weight-loss drugs for diabetes, we assume that they will avail diabetes drugs, while eligible patients without diabetes will access weight-loss drugs as obesity drugs. Thus, the proportions of individuals in BMI category $i$ eligible for diabetes and obesity drugs are

$$e_i^d = \begin{cases} 0 & \text{if } i \in \{1,2\}, \\ d_i & \text{if } i \in \{3,4,5,6,7\}, \end{cases}$$

and

$$e_i^o = \begin{cases} 0 & \text{if } i \in \{1,2,3,4\}, \\ 1-d_i & \text{if } i \in \{5,6,7\}, \end{cases}$$

respectively.

**B. Diabetes prevalence among individuals in different BMI categories.** Denoting the diabetes prevalence among adults as $\mathcal{D}$ and the proportion of the population in BMI category $i$ among diabetic patients as $v_i$, diabetes prevalence among the population in BMI category $i$ is given by

$$d_i = \frac{\mathcal{D}Pv_i}{O_iP} = \frac{\mathcal{D}v_i}{O_i}. \quad [13]$$

The national diabetes statistics report (6) provides the diabetes prevalence among adults aged 18 years and older and the proportions of the population with a BMI of $25.0-30.0$ (overweight-I and overweight-II; i.e., $v_3+v_4$), $30.0-40.0$ (obesity-I and obesity-II; i.e., $v_5+v_6$), and $\geq 40$ (obesity-III; i.e., $v_7$) among adults with diabetes. We assume $v_3 = (v_3+v_4)\frac{O_3}{O_3+O_4}$, $v_4 = (v_3+v_4)\frac{O_4}{O_3+O_4}$, $v_5 = (v_5+v_6)\frac{O_5}{O_5+O_6}$, and $v_6 = (v_5+v_6)\frac{O_6}{O_5+O_6}$. Thus, we have

$$d_3 = \mathcal{D}\frac{v_3}{O_3} = \mathcal{D}\frac{v_3+v_4}{O_3}\frac{O_3}{O_3+O_4} = \mathcal{D}\frac{v_3+v_4}{O_3+O_4}, \quad [14]$$

$$d_4 = \mathcal{D}\frac{v_4}{O_4} = \mathcal{D}\frac{v_3+v_4}{O_4}\frac{O_4}{O_3+O_4} = \mathcal{D}\frac{v_3+v_4}{O_3+O_4}, \quad [15]$$

$$d_5 = \mathcal{D}\frac{v_5}{O_5} = \mathcal{D}\frac{v_5+v_6}{O_5}\frac{O_5}{O_5+O_6} = \mathcal{D}\frac{v_5+v_6}{O_5+O_6}, \quad [16]$$

$$d_6 = \mathcal{D}\frac{v_6}{O_6} = \mathcal{D}\frac{v_5+v_6}{O_6}\frac{O_6}{O_5+O_6} = \mathcal{D}\frac{v_5+v_6}{O_5+O_6}, \quad [17]$$

and

$$d_7 = \frac{\mathcal{D}v_7}{O_7}. \quad [18]$$

### 5. Individuals' willingness and adherence to take weight-loss drugs

According to a KFF Health Tracking Poll, of those who are overweight or obese, 67% are very or somewhat interested in taking weight-loss drugs, while 8% are currently taking these drugs (7). Therefore, we assume that 75% of eligible individuals are willing to take weight-loss drugs in our base case scenario. While 11% of participants in the Health Tracking Poll are strongly against taking these drugs, 14% are only somewhat disinclined. To consider an optimistic scenario, we assume that those who are somewhat not interested would also be willing to take the drugs and thereby specify the willingness to take weight-loss drugs to be 89%.

Additionally, we assume that weight loss is only experienced among individuals with a medication adherence percentage greater than 80% for a year. Previous studies reported adherence rates of 27.2% (8) and 48.85% (9) among individuals without and with diabetes, respectively. In the optimistic scenario, we assume a perfect adherence.

We denote the willingness to take weight-loss drugs among the eligible population as $\alpha$ and the adherence rates among obesity and diabetes drug users as $\beta_o$ and $\beta_d$, respectively. Therefore, for our two scenarios, we set the parameters as

- Base case scenario: $\alpha = 0.75$, $\beta_o = 0.272$, and $\beta_d = 0.4885$;
- Optimistic scenario: $\alpha = 0.89$, $\beta_o = 1$, and $\beta_d = 1$.

### 6. Weight loss resulting from drug use

Data from a clinical trial of semaglutide provided (i) the mean weight loss percentage in people without type 2 diabetes (16.9%), (ii) the percentages of participants with a weight loss percentage greater than 5% (92.4%), 10% (74.8%), 15% (54.8%), and 20% (34.8%), and (iii) the maximum (50%) and minimum (-10%) weight loss percentages (10). We fitted a Beta distribution to the normalized data to construct a distribution of weight loss experienced by individuals without diabetes using weight-loss drugs.

A previous study (11) found that patients with type 2 diabetes had a lower percentage of weight loss compared with those without type 2 diabetes. Thus, we assume that the weight loss percentage is $\gamma\eta$ for patients with diabetes, where $\gamma$ is the weight loss percentage for patients without diabetes and $\eta$ is the ratio between weight loss percentages among patients with and without diabetes. We set $\eta \in [0.610, 0.619]$ based on results in (11).

### 7. Annual deaths with access to weight-loss drugs

To assess the impact of accessing weight-loss drugs, we calculate the expected number of annual deaths under two scenarios of accessibility to weight-loss drugs among those eligible:

1. Current uptake level,
2. Expanded access (full access to all eligible population).

From each population-level sample, we first sample individuals taking and adherent to obesity and diabetes drugs and then recalculate their BMI after taking drugs. If the recalculated BMI values fall into a different BMI category, the individuals are moved accordingly, and the proportion of the total population in each BMI category is recalculated.

**A. Sampling individuals taking and adherent to weight-loss drugs in different BMI categories.** The proportion of eligible individuals who take the drugs is dependent on the access and willingness ($\alpha$; section 5) to take the drugs. In the current uptake scenario, willingness is implicitly accounted for. The quantity of weight loss experienced is dependent on their adherence rate ($\beta_o$ and $\beta_d$; section 5) and whether they have type 2 diabetes or not.

Depending on the proportion of eligible individuals in each BMI category ($e_i^o O_i P$ and $e_i^d O_i P$) accessing weight-loss drugs, we sample the numbers of individuals in BMI category $i$ taking and adherent to obesity ($\beta_o r^o e_i^o O_i P$) and diabetes ($\beta_d r^d e_i^d O_i P$) drugs, where $r^o$ and $r^d$ are the uptake rates of obesity and diabetes drugs among the eligible population, respectively. We denote $r^o = r^o_{current}$ and $r^d = r^d_{current}$ under the current uptake scenario and $r^d = r^o = \alpha$ under the expanded access scenario. The sets of individuals taking diabetes and obesity drugs and expected to lose weight are denoted as $\mathcal{S}_D$ and $\mathcal{S}_O$, respectively. We have $|\mathcal{S}_D| = \sum_i \beta_d r^d e_i^d O_i P$ and $|\mathcal{S}_O| = \sum_i \beta_o r^o e_i^o O_i P$.

According to a study in 2021, the percentage of patients with type 2 diabetes treated with a GLP-1 receptor agonist was 10.7% (12). Another study in 2022 reported that 13.6% of individuals with type 2 diabetes were prescribed GLP-1 receptor agonists (13). Therefore, we assume that $r^d_{current} \in [0.107, 0.136]$. We use data from a recent KFF Health Tracking Poll (14) to obtain the current uptake rate of weight-loss drugs among those eligible solely for obesity drugs ($r^o_{current} = 0.108$).

**B. Redistribution of the population across BMI categories resulting from drug use.** For an overweight or obese individual $s$ with weight $W_s$ (kg) and height $H_s$ (m), the BMI value without taking weight-loss drugs is

$$BMI_s = W_s/H_s^2. \quad [19]$$

For those taking obesity drugs, the recalculated BMI value is

$$BMI'_s = W_s(1-\gamma)/H_s^2, \quad [20]$$

and for those taking diabetes drugs, the value is

$$BMI'_s = W_s(1-\gamma\eta)/H_s^2. \quad [21]$$

Thus, for given sets of individuals taking weight-loss drugs ($\mathcal{S}_D$ and $\mathcal{S}_O$), the recalculated percentage of individuals in BMI category $i$ is

$$O'_i(\mathcal{S}_D, \mathcal{S}_O) = \frac{\sum_s \mathcal{I}(\mathcal{B}_i^{min} \leq BMI'_s < \mathcal{B}_i^{max} | \mathcal{S}_D, \mathcal{S}_O)}{N_{pop}}, \quad [22]$$

where $\mathcal{I}(\cdot)$ is an identity function.

Therefore, the number of individuals moving from BMI category $i$ to $j$ resulting from drug use is

$$X_{ij}(\mathcal{S}_D, \mathcal{S}_O) = \sum_s \mathcal{I}(\mathcal{B}_i^{min} \leq BMI_s < \mathcal{B}_i^{max}, \mathcal{B}_j^{min} \leq BMI'_s < \mathcal{B}_j^{max} | \mathcal{S}_D, \mathcal{S}_O), \quad [23]$$

and the fraction of individuals moving from BMI category $i$ to $j$ resulting from drug use is

$$M_{ij}(\mathcal{S}_D, \mathcal{S}_O) = \frac{X_{ij}(\mathcal{S}_D, \mathcal{S}_O)}{O_i N_{pop}}. \quad [24]$$

**C. Updated annual mortality resulting from drug use.** The updated annual mortality is

$$\mathcal{E}(\mathcal{S}_D, \mathcal{S}_O) = \sum_{i=1}^{n} \mu_i O'_i(\mathcal{S}_D, \mathcal{S}_O) P = \mu_2 P \sum_{i=1}^{n} \lambda_i O'_i(\mathcal{S}_D, \mathcal{S}_O). \quad [25]$$

Thus, the reduction in annual mortality compared to no drug access is

$$\overline{\mu} P - \mathcal{E}(\mathcal{S}_D, \mathcal{S}_O). \quad [26]$$

The updated annual mortality in age group $a$ is

$$\mathcal{E}^a(\mathcal{S}_D, \mathcal{S}_O) = \sum_{i=1}^{n} \mu_i O'_{i,a}(\mathcal{S}_D, \mathcal{S}_O) p_a P = \mu_2 p_a P \sum_{i=1}^{n} \lambda_i O'_{i,a}(\mathcal{S}_D, \mathcal{S}_O), \quad [27]$$

where $O'_{i,a}(\mathcal{S}_D, \mathcal{S}_O)$ is the recalculated proportion of individuals in BMI category $i$ among individuals in age group $a$ resulting from drug use. Thus, the reduction in annual mortality in age group $a$ compared to no drug access is

$$\overline{\mu} p_a P - \mathcal{E}^a(\mathcal{S}_D, \mathcal{S}_O). \quad [28]$$

The reduction in annual mortality attributed to diabetes drugs is

$$\Delta\mathcal{E}_D(\mathcal{S}_D, \mathcal{S}_O) = \frac{P}{N_{pop}} \sum_{s \in \mathcal{S}_D} \sum_{i=1}^{n} \sum_{j=1}^{n} (\mu_i - \mu_j) \mathcal{I}(\mathcal{B}_i^{min} \leq BMI_s < \mathcal{B}_i^{max}, \mathcal{B}_j^{min} \leq BMI'_s < \mathcal{B}_j^{max} | \mathcal{S}_D, \mathcal{S}_O). \quad [29]$$

The reduction attributed to diabetes drugs among age group $a$ is

$$\Delta\mathcal{E}_D^a(\mathcal{S}_D, \mathcal{S}_O) = \frac{p_a P}{|\mathcal{S}_a|} \sum_{s \in \mathcal{S}_D \cap \mathcal{S}_a} \sum_{i=1}^{n} \sum_{j=1}^{n} (\mu_i - \mu_j) \mathcal{I}(\mathcal{B}_i^{min} \leq BMI_s < \mathcal{B}_i^{max}, \mathcal{B}_j^{min} \leq BMI'_s < \mathcal{B}_j^{max} | \mathcal{S}_D, \mathcal{S}_O), \quad [30]$$

where $\mathcal{S}_a$ is the set of individuals in age group $a$. Then, the reduction among adults aged 18 to 64 and 65 and over are

$$\Delta\mathcal{E}_D^{18-64}(\mathcal{S}_D, \mathcal{S}_O) = \sum_{a \in \{18,19,20-29,30-39,40-49,50-59\}} \Delta\mathcal{E}_D^a(\mathcal{S}_D, \mathcal{S}_O) + \Delta\mathcal{E}_D^{60-69}(\mathcal{S}_D, \mathcal{S}_O)/2, \quad [31]$$

and

$$\Delta\mathcal{E}_D^{65+}(\mathcal{S}_D, \mathcal{S}_O) = \sum_{a \in \{70-79,80+\}} \Delta\mathcal{E}_D^a(\mathcal{S}_D, \mathcal{S}_O) + \Delta\mathcal{E}_D^{60-69}(\mathcal{S}_D, \mathcal{S}_O)/2, \quad [32]$$

respectively.

Similarly, the reduction in annual mortality attributed to obesity drugs is

$$\Delta\mathcal{E}_O(\mathcal{S}_D, \mathcal{S}_O) = \frac{P}{N_{pop}} \sum_{s \in \mathcal{S}_O} \sum_{i=1}^{n} \sum_{j=1}^{n} (\mu_i - \mu_j) \mathcal{I}(\mathcal{B}_i^{min} \leq BMI_s < \mathcal{B}_i^{max}, \mathcal{B}_j^{min} \leq BMI'_s < \mathcal{B}_j^{max} | \mathcal{S}_D, \mathcal{S}_O), \quad [33]$$

and the corresponding reduction in age group $a$ is

$$\Delta\mathcal{E}_O^a(\mathcal{S}_D,\mathcal{S}_O)=\frac{p_aP}{|\mathcal{S}_a|}\sum_{s\in\mathcal{S}_O\cap\mathcal{S}_a}\sum_{i=1}^{n}\sum_{j=1}^{n}(\mu_i-\mu_j)\mathcal{I}(\mathcal{B}_i^{min}\leq BMI_s<\mathcal{B}_i^{max},\mathcal{B}_j^{min}\leq BMI_s'<\mathcal{B}_j^{max}|\mathcal{S}_D,\mathcal{S}_O). \quad [34]$$

Then, the reduction among adults aged 18 to 64 and 65 and over are

$$\Delta\mathcal{E}_O^{18-64}(\mathcal{S}_D,\mathcal{S}_O)=\sum_{a\in\{18,19,20-29,30-39,40-49,50-59\}}\Delta\mathcal{E}_O^a(\mathcal{S}_D,\mathcal{S}_O)+\Delta\mathcal{E}_O^{60-69}(\mathcal{S}_D,\mathcal{S}_O)/2, \quad [35]$$

and

$$\Delta\mathcal{E}_O^{65+}(\mathcal{S}_D,\mathcal{S}_O)=\sum_{a\in\{70-79,80+\}}\Delta\mathcal{E}_O^a(\mathcal{S}_D,\mathcal{S}_O)+\Delta\mathcal{E}_O^{60-69}(\mathcal{S}_D,\mathcal{S}_O)/2, \quad [36]$$

respectively. Note that,

$$\Delta\mathcal{E}_D(\mathcal{S}_D,\mathcal{S}_O)+\Delta\mathcal{E}_O(\mathcal{S}_D,\mathcal{S}_O)=\overline{\mu}P-\mathcal{E}(\mathcal{S}_D,\mathcal{S}_O), \quad [37]$$

and

$$\Delta\mathcal{E}_D^a(\mathcal{S}_D,\mathcal{S}_O)+\Delta\mathcal{E}_O^a(\mathcal{S}_D,\mathcal{S}_O)=\overline{\mu}p_aP-\mathcal{E}^a(\mathcal{S}_D,\mathcal{S}_O). \quad [38]$$

### 8. Deaths averted by insurance categories and US states

We distribute the national estimates of averted deaths across all insurance categories and US states, using the prevalence of obesity and diabetes and drug accessibility as weights.

**A. Individuals eligible for weight-loss drugs in different insurance categories and US states.** For a specific insurance category/state $x$, the proportions of individuals eligible for diabetes and obesity drugs are $e_x^d$ and $e_x^o$, respectively. Here,

$$e_x^d=\sum_{i=3}^{7}d_i^xO_i^x, \quad [39]$$

and

$$e_x^o=\sum_{i=5}^{7}(1-d_i^x)O_i^x, \quad [40]$$

where $O_i^x$ and $d_i^x$ are the proportion of individuals in BMI category $i$ among those in insurance category/state $x$ and the proportion of diabetes patients among those in BMI category $i$, respectively. We calculate $d_i^x$ as

$$d_i^x=\frac{\mathcal{D}_xP_xv_i^x}{O_i^xP_x}=\frac{\mathcal{D}_xv_i^x}{O_i^x}, \quad [41]$$

where $P_x$ and $\mathcal{D}_x$ are the population size and diabetes prevalence in insurance category/state $x$. Also, $v_i^x$ is the proportion of diabetic individuals in BMI category $i$ in insurance category/state $x$. Inserting Eq. (41) into Eqs. (39) and (40), we have

$$e_x^d=\sum_{i=3}^{7}\frac{\mathcal{D}_xv_i^x}{O_i^x}O_i^x=\mathcal{D}_x\sum_{i=3}^{7}v_i^x, \quad [42]$$

and

$$e_x^o=\sum_{i=5}^{7}(1-\frac{\mathcal{D}_xv_i^x}{O_i^x})O_i^x=\sum_{i=5}^{7}O_i^x-\mathcal{D}_x\sum_{i=5}^{7}v_i^x. \quad [43]$$

Thus, the proportion of individuals in insurance category/state $x$ eligible for weight-loss drugs is

$$e_x=e_x^d+e_x^o=\mathcal{D}_x\sum_{i=3}^{4}v_i^x+\sum_{i=5}^{7}O_i^x. \quad [44]$$

Based on Eqs. (42)-(44), the prevalence of diabetes ($\mathcal{D}_x$), obesity ($\sum_{i=5}^{7}O_i^x$), overweight ($\sum_{i=3}^{4}v_i^x$)/obesity ($\sum_{i=5}^{7}v_i^x$) among diabetic patients for each insurance category/state are needed to calculate the proportion of individuals eligible for weight-loss drugs.

To obtain $e_x^d$ and $e_x^o$ for different insurance categories, we use national-level diabetes (6) and obesity prevalence (3) along with the prevalence of diabetes and obesity among other insurance categories compared to that among individuals with private insurance (15, 16). According to previous studies, Medicaid beneficiaries were 27% more likely to have obesity than those with private insurance (15), while the obesity prevalence is 14.7% lower in the uninsured than the insured (16). We assume that diabetes prevalence and overweight and obesity prevalence among diabetic patients in each insurance category are the same as those at the national level. We integrate these differential risks across insurance categories with the overall diabetes and obesity prevalence to obtain $e_x^d$ and $e_x^o$ for different insurance categories.

To obtain $e_x^d$ and $e_x^o$ for different states, we use state-level obesity prevalence from KFF analysis of CDC's 2021–2022 Behavioral Risk Factor Surveillance System (17), state-level diabetes prevalence (18), and overweight and obesity prevalence among diabetic patients from CDC's United States Diabetes Surveillance System (18). We normalize the state-level prevalence of obesity, diabetes, and overweight/obesity among diabetic patients to align with national-level data.

**B. Distribution of annual averted deaths across insurance categories/US states.** Denote diabetes and obesity drug accessibility for insurance category/state $x$ as $\mathcal{A}_x^d \in [0,1]$ and $\mathcal{A}_x^o \in [0,1]$, respectively. Under the current uptake scenario, uninsured people are unlikely to access high-cost weight-loss drugs, therefore, we assume

$$\mathcal{A}_{\text{Uninsured}}^d = \mathcal{A}_{\text{Uninsured}}^o = 0.$$

Since drugs for type 2 diabetes are covered by all other insurance categories, we assume

$$\mathcal{A}_{\text{Medicare}}^d = \mathcal{A}_{\text{Medicaid}}^d = \mathcal{A}_{\text{Private}}^d = 1.$$

Currently, approximately 50 million US adults have coverage for obesity drugs, with 40 million through commercial insurance and 10 million through Medicaid (19), thus we assume, under the current uptake scenario,

$$\Delta\mathcal{E}_O^{\text{Private}}(\mathcal{S}_D, \mathcal{S}_O) = 4\Delta\mathcal{E}_O^{\text{Medicaid}}(\mathcal{S}_D, \mathcal{S}_O),$$

where $\Delta\mathcal{E}_O^x(\mathcal{S}_D, \mathcal{S}_O)$ is the reduction in annual mortality attributed to obesity drugs in insurance category $x$. Additionally, 4.7% of current obesity drug users are aged 65 and older (14), thus, we have

$$\Delta\mathcal{E}_O^{\text{Medicare}}(\mathcal{S}_D, \mathcal{S}_O) = \Delta\mathcal{E}_O^{65+}(\mathcal{S}_D, \mathcal{S}_O) = 0.047\Delta\mathcal{E}_O(\mathcal{S}_D, \mathcal{S}_O).$$

Under the expanded access scenario, for all insurance categories,

$$\mathcal{A}_x^d = \mathcal{A}_x^o = 1.$$

For all US states, $\mathcal{A}_x^d = \mathcal{A}_x^o = 1$ under both current uptake and expanded access scenarios. Then, the reduction in annual mortality attributed to diabetes and obesity drugs in insurance category/state $x$ is calculated as

$$\Delta\mathcal{E}_D^x(\mathcal{S}_D, \mathcal{S}_O) = \Delta\mathcal{E}_D(\mathcal{S}_D, \mathcal{S}_O)\frac{\mathcal{A}_x^d e_x^d P_x}{\sum_y \mathcal{A}_y^d e_y^d P_y},$$

and

$$\Delta\mathcal{E}_O^x(\mathcal{S}_D, \mathcal{S}_O) = \Delta\mathcal{E}_O(\mathcal{S}_D, \mathcal{S}_O)\frac{\mathcal{A}_x^o e_x^o P_x}{\sum_y \mathcal{A}_y^o e_y^o P_y},$$

respectively.

**Table S1. US adults by BMI category and their hazard ratios for all-cause mortality and share of annual mortality**

| Index ($i$) | BMI category | BMI range ($\mathcal{B}_i^{min}$–$\mathcal{B}_i^{max}$) | Prevalence ($O_i$) | Hazard ratio ($\lambda_i$) | Share of annual mortality |
|---|---|---|---|---|---|
| 1 | Underweight | $< 18.5$ | 1.13% | 1.894 | 1.87% |
| 2 | Normal | 18.5–25.0 | 27.71% | 1.000 | 24.18% |
| 3 | Overweight-I | 25.0–27.5 | 15.80% | 0.938 | 12.93% |
| 4 | Overweight-II | 27.5–30.0 | 14.40% | 1.002 | 12.59% |
| 5 | Obesity-I | 30.0–35.0 | 22.30% | 1.133 | 22.05% |
| 6 | Obesity-II | 35.0–40.0 | 10.74% | 1.445 | 13.54% |
| 7 | Obesity-III | $\geq 40.0$ | 7.92% | 1.856 | 12.83% |

**Table S2. US population aged 18 and older by age and gender ($p_{g,a}$, $p_a$)**

| Age | Gender | % Population | % Population (Monte Carlo samples) |
|---|---|---|---|
| 18 years | | 1.68 | |
| 18 years | Female | 0.82 | 0.82 |
| 18 years | Male | 0.86 | 0.86 |
| 19 years | | 1.65 | |
| 19 years | Female | 0.81 | 0.81 |
| 19 years | Male | 0.85 | 0.85 |
| 20–29 | | 16.85 | |
| 20–29 | Female | 8.27 | 8.26 |
| 20–29 | Male | 8.59 | 8.59 |
| 30–39 | | 17.60 | |
| 30–39 | Female | 8.72 | 8.72 |
| 30–39 | Male | 8.88 | 8.88 |
| 40–49 | | 15.82 | |
| 40–49 | Female | 7.90 | 7.89 |
| 40–49 | Male | 7.92 | 7.92 |
| 50–59 | | 16.12 | |
| 50–59 | Female | 8.13 | 8.13 |
| 50–59 | Male | 8.00 | 8.00 |
| 60–69 | | 15.39 | |
| 60–69 | Female | 7.99 | 7.98 |
| 60–69 | Male | 7.41 | 7.41 |
| 70–79 | | 10.04 | |
| 70–79 | Female | 5.43 | 5.43 |
| 70–79 | Male | 4.60 | 4.60 |
| 80 and over | | 4.84 | |
| 80 and over | Female | 2.94 | 2.93 |
| 80 and over | Male | 1.91 | 1.91 |

**Table S3. Proportion of the population eligible for weight-loss drugs and annual averted deaths by states under the expanded access compared to current uptake of weight-loss drugs.**

| State | Eligible (%) | Total deaths | Total deaths (obesity) | Total deaths (diabetes) |
|---|---|---|---|---|
| Rhode Island | 41.41 | 132 | 97 | 35 |
| Maryland | 44.85 | 809 | 609 | 200 |
| Alabama | 54.34 | 801 | 545 | 256 |
| Oklahoma | 54.72 | 642 | 465 | 177 |
| Pennsylvania | 44.31 | 1680 | 1247 | 433 |
| Kansas | 48.68 | 418 | 306 | 112 |
| South Carolina | 48.08 | 738 | 499 | 239 |
| Wisconsin | 47.98 | 831 | 664 | 167 |
| Tennessee | 52.43 | 1076 | 745 | 331 |
| Montana | 40.21 | 132 | 101 | 31 |
| Oregon | 42.94 | 533 | 401 | 132 |
| Illinois | 45.46 | 1669 | 1211 | 458 |
| Ohio | 50.96 | 1749 | 1277 | 473 |
| Connecticut | 40.61 | 429 | 304 | 125 |
| Missouri | 48.02 | 867 | 646 | 221 |
| South Dakota | 49.27 | 131 | 99 | 33 |
| Nevada | 44.76 | 414 | 294 | 120 |
| Idaho | 45.09 | 256 | 199 | 57 |
| Indiana | 51.26 | 1023 | 754 | 269 |
| New Mexico | 43.51 | 267 | 177 | 90 |
| Minnesota | 45.38 | 762 | 598 | 164 |
| Georgia | 47.49 | 1510 | 1072 | 438 |
| New York | 42.09 | 2412 | 1703 | 710 |
| Virginia | 46.41 | 1176 | 851 | 325 |
| California | 39.77 | 4504 | 2997 | 1507 |
| Iowa | 50.70 | 477 | 377 | 100 |
| Utah | 42.79 | 424 | 331 | 93 |
| Massachusetts | 38.02 | 778 | 594 | 184 |
| Nebraska | 48.62 | 280 | 216 | 65 |
| Florida | 39.12 | 2540 | 1841 | 699 |
| New Jersey | 40.94 | 1107 | 809 | 298 |
| Colorado | 34.15 | 584 | 448 | 136 |
| North Carolina | 45.43 | 1416 | 1005 | 412 |
| Delaware | 51.48 | 153 | 113 | 40 |
| Kentucky | 49.69 | 652 | 447 | 205 |
| Mississippi | 55.90 | 478 | 331 | 147 |
| North Dakota | 48.36 | 110 | 84 | 26 |
| Alaska | 41.94 | 90 | 71 | 19 |
| West Virginia | 56.08 | 290 | 207 | 83 |
| Washington | 42.79 | 976 | 747 | 229 |
| Arkansas | 50.32 | 447 | 317 | 129 |
| Hawaii | 34.54 | 145 | 107 | 38 |
| Arizona | 45.01 | 967 | 708 | 260 |
| Maine | 43.11 | 175 | 136 | 39 |
| New Hampshire | 40.16 | 165 | 130 | 34 |
| Michigan | 46.81 | 1377 | 1059 | 318 |
| Vermont | 34.59 | 66 | 49 | 16 |
| Louisiana | 52.48 | 702 | 502 | 201 |
| Texas | 48.10 | 4210 | 2996 | 1214 |
| District of Columbia | 34.35 | 67 | 46 | 21 |
| Wyoming | 44.98 | 77 | 62 | 15 |

**Table S4. Redistribution of individuals in a BMI category under the current uptake of weight-loss drugs**

| BMI category | Underweight (%) | Normal (%) | Overweight-I (%) | Overweight-II (%) | Obesity-I (%) | Obesity-II (%) | Obesity-III (%) |
|---|---|---|---|---|---|---|---|
| Overweight | 0 | 0.48 | 51.51 | 48.02 | 0 | 0 | 0 |
| Overweight-I | 0 | 0.67 | 99.33 | 0 | 0 | 0 | 0 |
| Overweight-II | 0 | 0.27 | 0.41 | 99.32 | 0 | 0 | 0 |
| Obesity | 0 | 0.4 | 0.6 | 0.78 | 51.4 | 28.84 | 17.98 |
| Obesity-I | 0 | 0.7 | 0.96 | 1.11 | 97.22 | 0.01 | 0 |
| Obesity-II | 0 | 0.11 | 0.3 | 0.62 | 1.9 | 97.06 | 0.01 |
| Obesity-III | 0 | 0 | 0.04 | 0.13 | 0.89 | 1.79 | 97.15 |

**Table S5. Redistribution of individuals in a BMI category under the expanded access of weight-loss drugs**

| BMI category | Underweight (%) | Normal (%) | Overweight-I (%) | Overweight-II (%) | Obesity-I (%) | Obesity-II (%) | Obesity-III (%) |
|---|---|---|---|---|---|---|---|
| Overweight | 0 | 2.92 | 50.76 | 46.31 | 0.01 | 0 | 0 |
| Overweight-I | 0 | 4.08 | 95.91 | 0.01 | 0 | 0 | 0 |
| Overweight-II | 0 | 1.68 | 2.53 | 95.78 | 0.01 | 0 | 0 |
| Obesity | 0.01 | 2.75 | 4.05 | 5.23 | 47.2 | 25.75 | 15.01 |
| Obesity-I | 0.02 | 4.83 | 6.49 | 7.31 | 81.3 | 0.05 | 0 |
| Obesity-II | 0 | 0.77 | 2.1 | 4.25 | 12.61 | 80.21 | 0.05 |
| Obesity-III | 0 | 0.03 | 0.27 | 0.92 | 6.04 | 11.72 | 81.02 |

**Table S6. Redistribution of individuals in a BMI category under the expanded access of weight-loss drugs and increased willingness and adherence rate to take weight-loss drugs**

| BMI category | Underweight (%) | Normal (%) | Overweight-I (%) | Overweight-II (%) | Obesity-I (%) | Obesity-II (%) | Obesity-III (%) |
|---|---|---|---|---|---|---|---|
| Overweight | 0 | 7.1 | 49.48 | 43.4 | 0.02 | 0 | 0 |
| Overweight-I | 0 | 9.93 | 90.03 | 0.03 | 0 | 0 | 0 |
| Overweight-II | 0 | 4.07 | 6.12 | 89.77 | 0.03 | 0 | 0 |
| Obesity | 0.05 | 11.79 | 15.91 | 19.36 | 32.44 | 14.55 | 5.89 |
| Obesity-I | 0.1 | 20.66 | 24.97 | 25.89 | 28.18 | 0.19 | 0 |
| Obesity-II | 0 | 3.37 | 9.12 | 17.45 | 45.43 | 24.43 | 0.2 |
| Obesity-III | 0 | 0.14 | 1.16 | 4.03 | 23.84 | 39.34 | 31.48 |

**Table S7. Model parameters**

| Parameter | Symbol | Value | Reference |
|---|---|---|---|
| $\lambda_i$ | The hazard ratio for all-cause mortality in BMI category $i$ | Table S1 | (2) |
| $O_i^{g,a}$ | Percentage of the population in BMI category $i$ among individuals in gender group $g$ and age group $a$ without access to weight-loss drugs | | (3) |
| $p_{g,a}$ | Proportion of the population in gender group $g$ and age group $a$ among all adults | Table S2 | (4) |
| $O_i^a$ or $O_i^x$ | Percentage of the population in BMI category $i$ among individuals in age group $a$ or insurance category/state $x$ without access to weight-loss drugs | | (3) |
| $p_a$ | Proportion of the population in age group $a$ among all adults | Table S2 | (4) |
| $O_i$ | Percentage of the population in BMI category $i$ among adults without access to weight-loss drugs | Table S1 | (3, 4) |
| $N_{pop}$ | Population-level sample size | 100,000 | |
| $P_{total}$ | Total population in the US | 333,287,562 | (20) |
| $p_{\geq 18}$ | Proportion of adults aged 18 years and older among US population | 78.01% | (4) |
| $P$ | Total adults population in the US | 260,004,955 | (4, 20) |
| $\overline{\mu}$ | Annual mortality rate without drug access | 0.009841 | (21) |
| $\mu_i$ | Annual mortality rate without drug access for BMI category $i$ | | |
| $e_i$ or $e_x$ | Proportion of individuals eligible for weight-loss drugs in BMI category $i$ or insurance category/state $x$ | | |
| $e_i^d$ or $e_x^d$ | Proportion of individuals eligible for diabetes drugs in BMI category $i$ or insurance category/state $x$ | | |
| $e_i^o$ or $e_x^o$ | Proportion of individuals eligible for obesity drugs in BMI category $i$ or insurance category/state $x$ | | |
| $d_i$ | Diabetes prevalence among the population in BMI category $i$ | | |
| $d_i^x$ | Diabetes prevalence among the population in BMI category $i$ and insurance category/state $x$ | | |
| $\mathcal{D}$ | Diabetes prevalence among adults | 13.2%–16.4% | (6) |
| $\mathcal{D}_x$ | Diabetes prevalence among adults in insurance category/state $x$ | | (6, 18) |
| $v_i$ | Proportion of individuals in BMI category $i$ among diabetic patients | | |
| $v_i^x$ | Proportion of individuals in BMI category $i$ among diabetic patients in insurance category/state $x$ | | |
| $v_3 + v_4$ | Proportion of the population in overweight-I and overweight-II categories among diabetic patients | 24.0%–30.0% | (6) |
| $v_5 + v_6$ | Proportion of the population in obesity-I and obesity-II categories among diabetic patients | 43.3%–51.0% | (6) |
| $v_7$ | Proportion of the population in obesity-III category among diabetic patients | 12.4%–19.7% | (6) |
| $\alpha$ | Willingness to take weight-loss drugs among the eligible population | 0.75; 0.89 | (7) |
| $\beta_o$ | Adherence rate among obesity drug users | 0.272; 1 | (8) |
| $\beta_d$ | Adherence rate among diabetes drug users | 0.4885; 1 | (9) |
| $\gamma$ | Weight loss percentage for patients without diabetes | | (10) |
| $\eta$ | Ratio between weight loss percentages among patients with and without diabetes | 61.0%–61.9% | (11) |
| $r^d$ | Diabetes drug uptake rate among the eligible population | | |
| $r^o$ | Obesity drug uptake rate among the eligible population | | |
| $r^d_{current}$ | Current diabetes drug uptake rate among the eligible population | 10.7%–13.6% | (12, 13) |
| $r^o_{current}$ | Current obesity drug uptake rate among the eligible population | 10.8% | (14) |
| $\mathcal{S}_D$ | Set of individuals taking diabetes drugs and expected to lose weight | | |
| $\mathcal{S}_O$ | Set of individuals taking obesity drugs and expected to lose weight | | |
| $\mathcal{S}_a$ | Set of individuals in age group $a$ in the population-level sample | | |
| $O_i'(\mathcal{S}_D, \mathcal{S}_O)$ | Updated percentage of the population in BMI category $i$ resulting from drug use | | |
| $X_{ij}(\mathcal{S}_D, \mathcal{S}_O)$ | Number of individuals moving from BMI category $i$ to $j$ resulting from drug use | | |
| $M_{ij}(\mathcal{S}_D, \mathcal{S}_O)$ | Fraction of individuals moving from BMI category $i$ to $j$ resulting from drug use | | |
| $\mathcal{E}(\mathcal{S}_D, \mathcal{S}_O)$ | Updated annual mortality | | |
| $\mathcal{E}^a(\mathcal{S}_D, \mathcal{S}_O)$ | Updated annual mortality in age group $a$ | | |
| $\Delta\mathcal{E}_D(\mathcal{S}_D, \mathcal{S}_O)$ | Reduction in annual mortality compared to no drug access attributed to diabetes drugs | | |
| $\Delta\mathcal{E}_D^a(\mathcal{S}_D, \mathcal{S}_O)$ | Reduction in annual mortality compared to no drug access attributed to diabetes drugs in age group $a$ | | |
| $\Delta\mathcal{E}_D^x(\mathcal{S}_D, \mathcal{S}_O)$ | Reduction in annual mortality compared to no drug access attributed to diabetes drugs in insurance category/state $x$ | | |
| $\Delta\mathcal{E}_O(\mathcal{S}_D, \mathcal{S}_O)$ | Reduction in annual mortality compared to no drug access attributed to obesity drugs | | |
| $\Delta\mathcal{E}_O^a(\mathcal{S}_D, \mathcal{S}_O)$ | Reduction in annual mortality compared to no drug access attributed to obesity drugs in age group $a$ | | |
| $\Delta\mathcal{E}_O^x(\mathcal{S}_D, \mathcal{S}_O)$ | Reduction in annual mortality compared to no drug access attributed to obesity drugs in insurance category/state $x$ | | |
| $P_x$ | Population size of insurance category/state $x$ | | |
| $\mathcal{A}_x^d$ | Diabetes drug accessibility for insurance category/state $x$ | | |
| $\mathcal{A}_x^o$ | Obesity drug accessibility for insurance category/state $x$ | | |

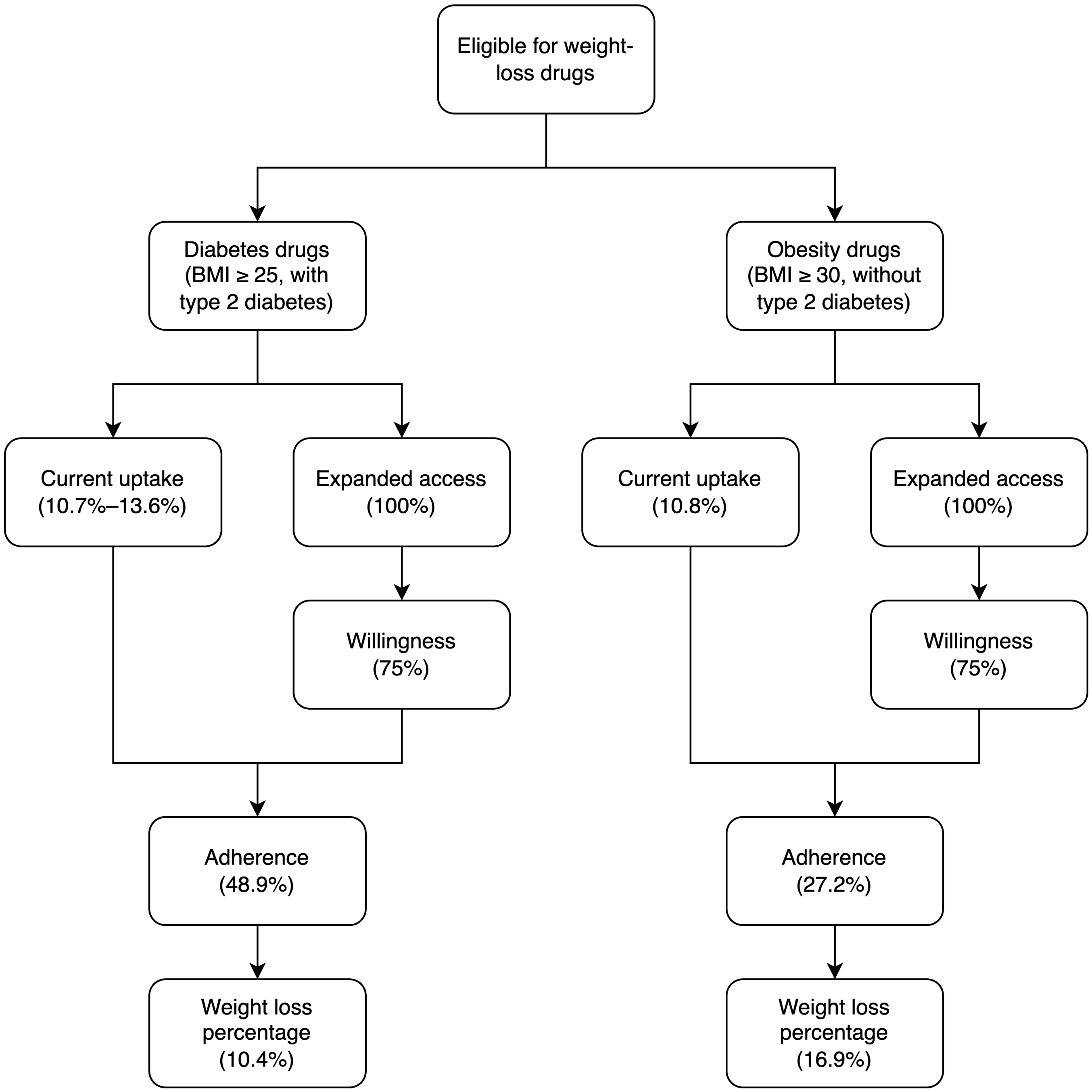


**Fig. S1.** Weight loss among eligible individuals under the current uptake level of and expanded access to weight-loss drugs.

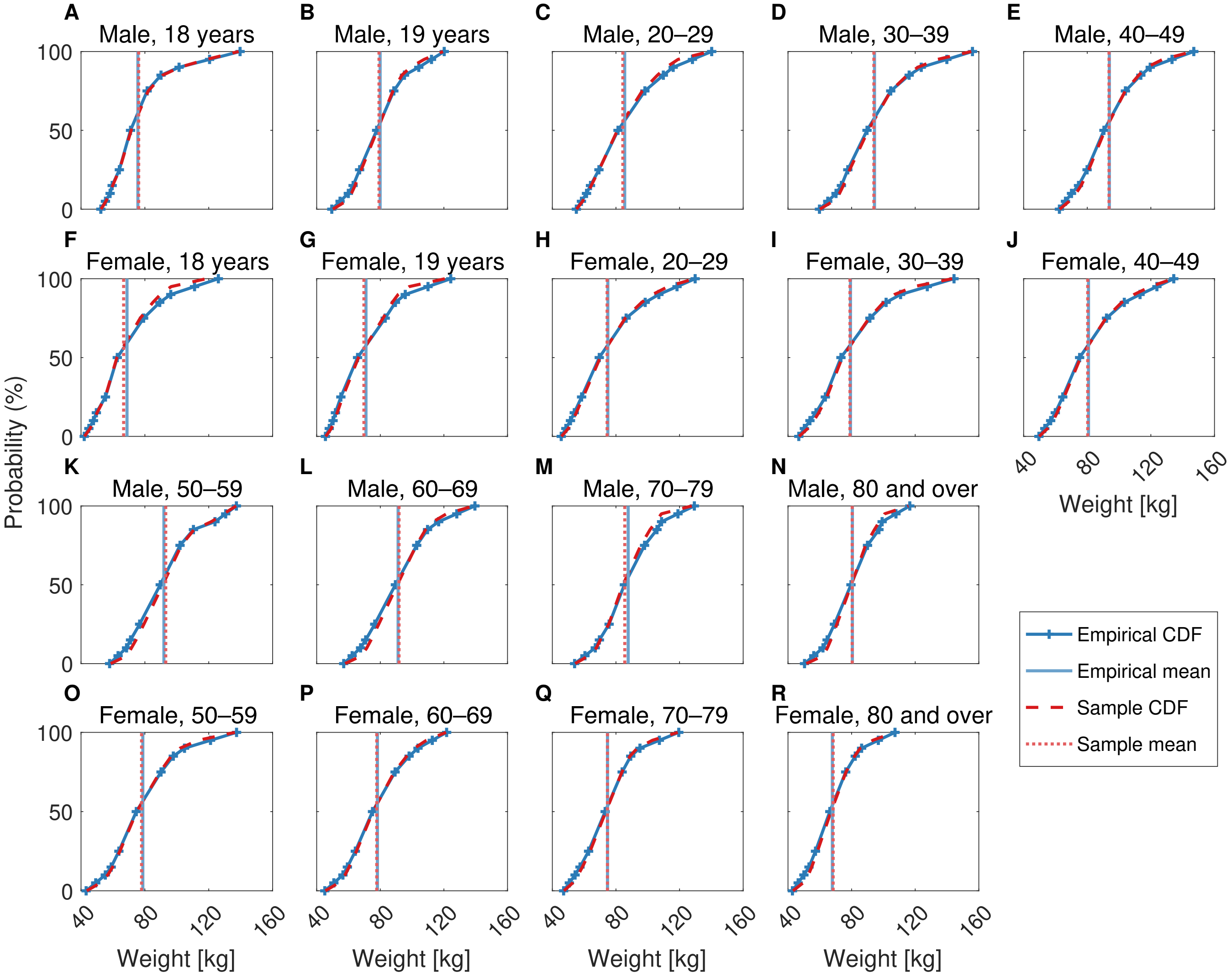


**Fig. S2.** Comparison of the cumulative distribution of weight between empirical (blue solid curves) and Monte Carlo sampling results (red dashed curves) for each gender-age group. Blue solid and red dotted vertical lines represent the mean of the empirical cumulative distribution and the cumulative distribution of Monte Carlo samples, respectively. The cumulative distribution of Monte Carlo samples is obtained by averaging 1000 Monte Carlo simulations.

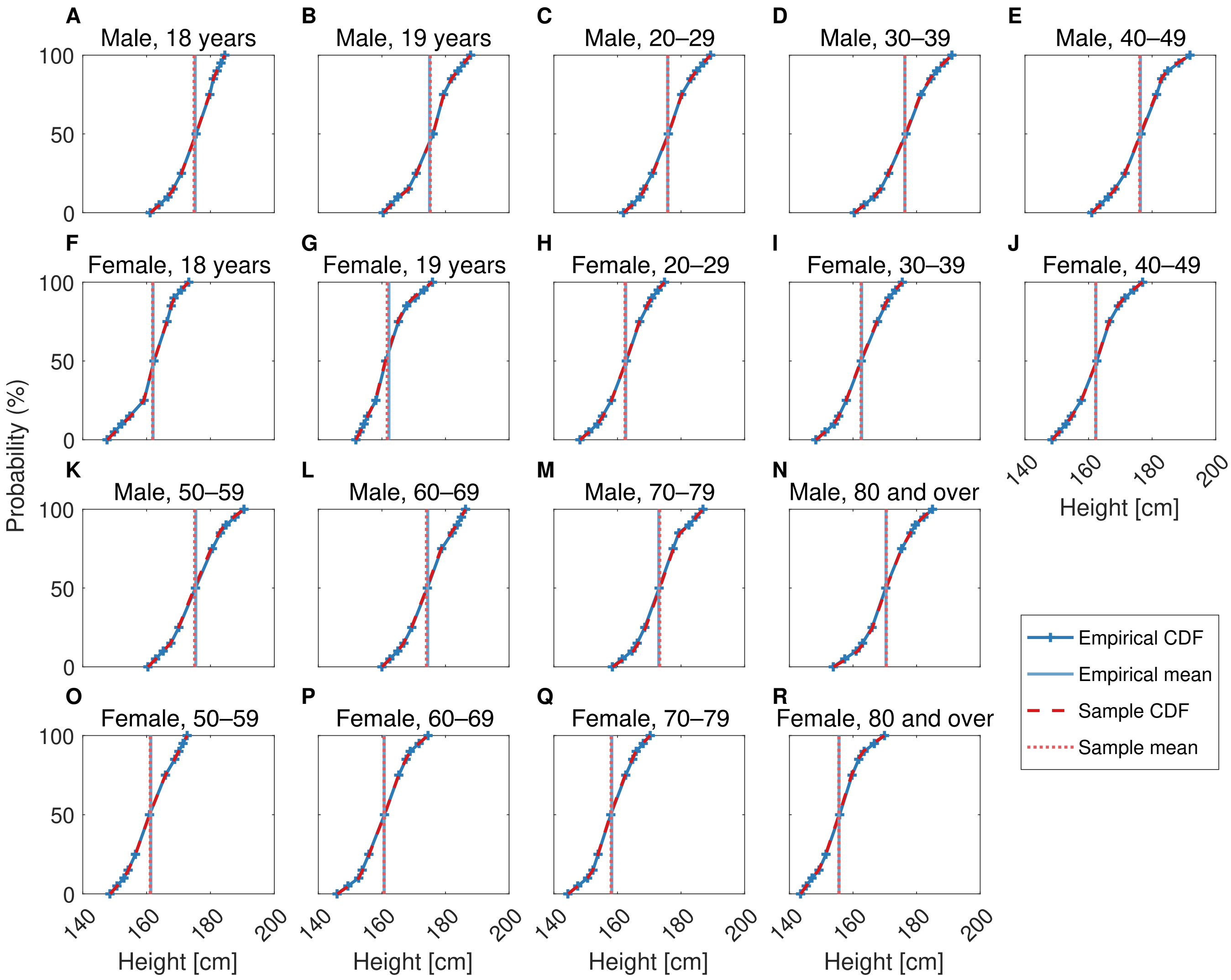


**Fig. S3.** Comparison of the cumulative distribution of height between empirical (blue solid curves) and Monte Carlo sampling results (red dashed curves) for each gender-age group. Blue solid and red dotted vertical lines represent the mean of the empirical cumulative distribution and the cumulative distribution of Monte Carlo samples, respectively. The cumulative distribution of Monte Carlo samples is obtained by averaging 1000 Monte Carlo simulations.

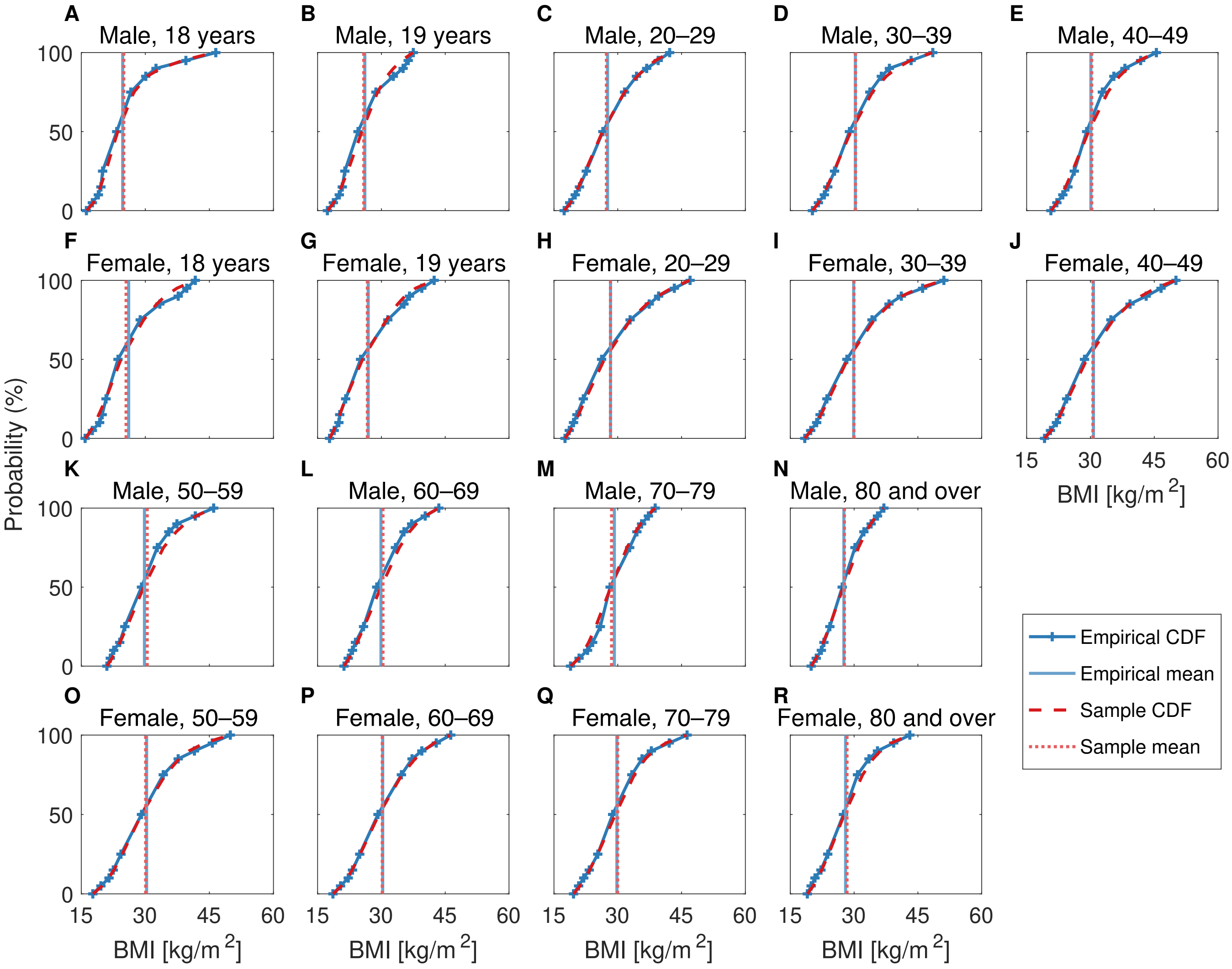


**Fig. S4.** Comparison of the cumulative distribution of BMI between empirical (blue solid curves) and Monte Carlo sampling results (red dashed curves) for each gender-age group. Blue solid and red dotted vertical lines represent the mean of the empirical cumulative distribution and the cumulative distribution of Monte Carlo samples, respectively. The cumulative distribution of Monte Carlo samples is obtained by averaging 1000 Monte Carlo simulations.
